# Socio-economic patterns of diet, obesity, and biomarkers for cardiovascular disease among Indian adolescents

**DOI:** 10.1101/2024.09.23.24314192

**Authors:** Neelam Kalita, Susan Griffin, Sumit Mazumdar

## Abstract

**Background:** The impact of socioeconomic (SES) factors, maternal education and household wealth on diet and consequently on a host of cardiovascular disease (CVD) biomarkers is rarely examined among Indian adolescents, who are in a critical development phase. This study examines the socio-economic patterning of dietary behaviour and its correlation with overweight/obesity and CVD biomarkers among Indian adolescents aged 10 to 19 years.

**Methods:** We use nationally representative data on dietary patterns and CVD biomarkers from the Comprehensive National Nutrition Dataset, a nationally representative survey of 35,830 adolescents conducted from 2016 to 2018. Dietary pattern is assessed using a summary indicator-Dietary Diversity Score based on 17 specific food groups. Overweight/obesity and the biomarkers, including the ratio of total cholesterol to high-density lipoprotein-cholesterol, serum triglycerides, hypertension, and pre-diabetic/diabetic, are analysed.

**Results:** Adolescents from higher SES have higher dietary diversity than those from lower SES. This socio-economic patterning is concentrated on higher daily consumption of certain foods such as fats and oil, sugar and jaggery and those high in fats, sugar, and salt (HFSS) comprising-junk food, fried food, sweets, and aerated drinks among those from high SES and urban areas. This, in turn, may explain the concentration of overweight/obesity in this cohort. The SES gradient in diet, overweight/obesity, and associated CVD biomarkers among Indian adolescents remains positive for the household wealth index (worse health for the more economically advantaged) and urban dwellers. However, it is starting to shift towards negative for maternal education, resulting in an inverted U shape (better health for the lowest and highest socially advantaged, worse health for those in the middle).

**Conclusions:** There is a clustering of overweight/obesity and CVD biomarkers among adolescents from urban and wealthier households that may be associated with increased consumption of HFSS in this cohort.

## Introduction

Cardiovascular diseases (CVDs) are a leading cause of disease burden and death in India, accounting for one-fifth of the global CVD related deaths.^1 2^ Major transitions in population-level changes across six areas-nutritional, epidemiological, demographic, environmental, socio-cultural and economic factors can largely explain India’s CVD epidemic.^3 4^ Focussing on nutrition, there has been a shift towards decreased consumption of healthy foods (e.g. coarse cereals, pulses, fruits and vegetables) and corresponding increased consumption of foods with high energy and high in fats, sugar and salt (HFSS), leading to obesity and nutrition-related non-communicable diseases (NCDs).^3 5-8^ Factors such as increased household income, reduced time for meal preparation, access to processed food and marketing of sugar sweetened beverages, poor awareness regarding healthy eating have resulted in traditional diets being replaced by prepared and/or processed foods that are HFSS.^3 9^ To address this growing trend, which is a major public health concern, epidemiological studies^1 2 5-11^ have emphasised on improving the overall diet quality to reduce CVD burden and related deaths.

Socioeconomic factors such as levels of -income and -education are closely related with diet quality. They impact health and contribute to the CVD burden by attributing to a host of risk factors-biological, behavioural, and psychological.^12 13^ Previous studies examining the relationship between socio-economic status (SES) and CVD biomarkers (including, unhealthy weight, hypertension, diabetes, lipid anomalies) in India have reported mixed findings: some found the prevalence of the biomarkers higher among low SES groups,^14-17^ others reported higher prevalence among those from high SES,^18 19^ whereas some reported no clear trends.^20 21^ Such contrasting findings have initiated researchers to hypothesize that the relationship between SES and CVD biomarkers follows a reversal trend over the course of a country’s economic progress: initially these factors are concentrated among high SES, as a country develops, they become more prevalent among low SES.^15 22 23^ Most of the existing literature focus on adults; the socio-economic patterning of dietary behaviour and their correlation with the prevalence of CVD biomarkers among adolescents are unclear. Therefore, we test this hypothesis in India’s adolescents by examining: i) the shape and nature of the SES gradients for dietary behaviour and CVD biomarkers; ii) how these gradients vary by gender and area of residence. We address this by using data from the Comprehensive National Nutrition Survey (CNNS) dataset.

Adolescence, between 10 and 19 years, is a critical period for developing later health and disease. With 253 million, India has the largest adolescent population worldwide and faces a double burden of undernutrition and overnutrition.^24-26^ To address this double burden, optimal public health interventions focusing on promoting healthy eating behaviours and other lifestyle choices in adolescence are required.

In India, nationally representative literature on CVD biomarkers among adolescents is scant. Previous studies assessing CVD biomarkers, based on sub-regional and smaller sub-groups of population, found substantial burden of lipid anomalies among adolescents.^27-29 30 31^ They mainly used healthcare sources for data sources. Furthermore, no nationally representative study collected data on their nutritional status, particularly for the young adolescents aged 10 to 11 years. In this study, we addressed these research gaps by using the CNNS dataset that collected nationally representative data across India, for the first time, on nutrition, socioeconomic variables, and the prevalence of CVD biomarkers for adolescents aged 10 to 19 years.^25 32^

## Subjects and Methods

This is a secondary analysis of the publicly released data from the CNNS in India, conducted between February 2016 and October 2018, by the Population Council, under the leadership of the Ministry of Health and Family Welfare and UNICEF. The CNNS collected for the first-time country-wide cross-sectional data on nutritional status of pre-schoolers (0 - 4 years), children of school-going age (5 – 9 years) and adolescents (10 – 19 years), anthropometric measures (height, weight, mid-upper arm circumference, triceps skinfold thickness, subscapular skinfold thickness, waist circumference), and biomarkers of NCDs (blood pressure, blood sugar, lipid profiles) in school-age children and adolescents. The survey excluded individuals with chronic illness, major physical deformity, cognitive disabilities, acute febrile infectious illness and pregnancy.^33^

Data on household characteristics, environmental conditions, individual dietary intake, health status and anthropometry were collected from 35,830 adolescents, aged 10 – 19 years. Of these, 18,425 were boys (51.42%) and 17,405 (48.58%) girls, respectively. For adolescents aged 10-14 years, a parent was asked to be present during the interview to help the adolescent respond. For all the adolescents (10-19 years), parents answered questions about parental- and household factors (e.g., education level, sanitation).

### Dietary data

Data on individual dietary intake was collected using Food Frequency Questionnaire that collected information on daily and seven-day consumption frequency of 17 food groups over the previous week: cereals, dairy, pulses or beans, roots or tubers, dark green leafy vegetables, other vegetables, fruits, eggs, fish, chicken or meat, nuts or seeds, fats or oils, sugar or jaggery, fried food, junk food, sweets, and aerated drinks. This information was used to measure a summary indicator: Dietary Diversity Score (DDS), a commonly used dietary measure showing dietary pattern in resource-constraint settings as the estimation is straightforward and do not require a food composition database.^34^

DDS, adapted and modified from the methods used in the existing literature^14 35-37^ and the Indian dietary guidelines,^10 38^ was calculated including based on the number of food groups consumed per week. Of the 17 food groups, 13 recommended by the Indian Council of Medical Research (ICMR)^10^ and Food and Agriculture Organization (FAO),^39^ was given a score of 1 for each intake and counted only once even if it was consumed more than once per week. For the remaining four HFSS food groups: fried food, junk food, sweets, and aerated drinks, the scoring was reversed to align with the guidance from the Indian Academy of Paediatrics Guidelines on the Fast and Junk Foods, Sugar Sweetened Beverages, Fruit Juices, and Energy Drinks.^38^ A score of 1 for no intake and 0 for intake was assigned and counted only once even if it was consumed more than once per week. The total score ranged from 0 to 17; 17 representing full compliance to dietary guidelines whereas 0 representing no compliance.

### CVD biomarkers

Five biomarkers are included:^40^

i. Obesity or overweight, measured by BMI for age Z-scores as:

- Overweight: >1 to 2 standard deviations (SD)
- Obesity: >2 SD.
ii. Two lipid anomalies that increase the risk for developing heart diseases:

- High ratio of Total cholesterol to High-Density Lipoprotein-cholesterol (HDL-c): >4.5
- High serum triglycerides:≥130 mg/dl
iii. Hypertension, measured by a systolic blood pressure level ≥140 mmHg, or a diastolic blood pressure level ≥ 90mmHg, respectively
iv. Pre-diabetic or diabetic status measured by glycosylated haemoglobin (HbA1c):

- Pre-diabetic: >5.6% and ≤6.4%
- Diabetic: >6.4%

### SES variables

Two primary indicators of SES, maternal educational attainment and household wealth index, are included.^13 41^ Levels of maternal education attainment were classified as: primary (up to 8 years); secondary (up to 12 years); graduate (up to 15 years); and higher (≥16 years). Wealth index was estimated based on a household’s possession of several common household items in the CNNS. Data on the household’s access to and type of water and sanitation facilities, source of lighting, type of cooking fuel, and ownership of assets such as house, land, livestock, television, radio, and vehicles was used. Households were given scores derived using principal component analysis adjusted for national and state-level weights. Wealth quintiles were computed by dividing the weighted distribution into five equal categories (viz, poorest (1), poor (2), middle (3), rich (4), richest (5)), each with 20% of the sample population.^40 42^

### Other characteristics

The dietary landscape and the prevalence of CVD biomarkers are explored for other indicators including area of residence, religion, and caste detailed in **Table 1**.

**Table 1.**
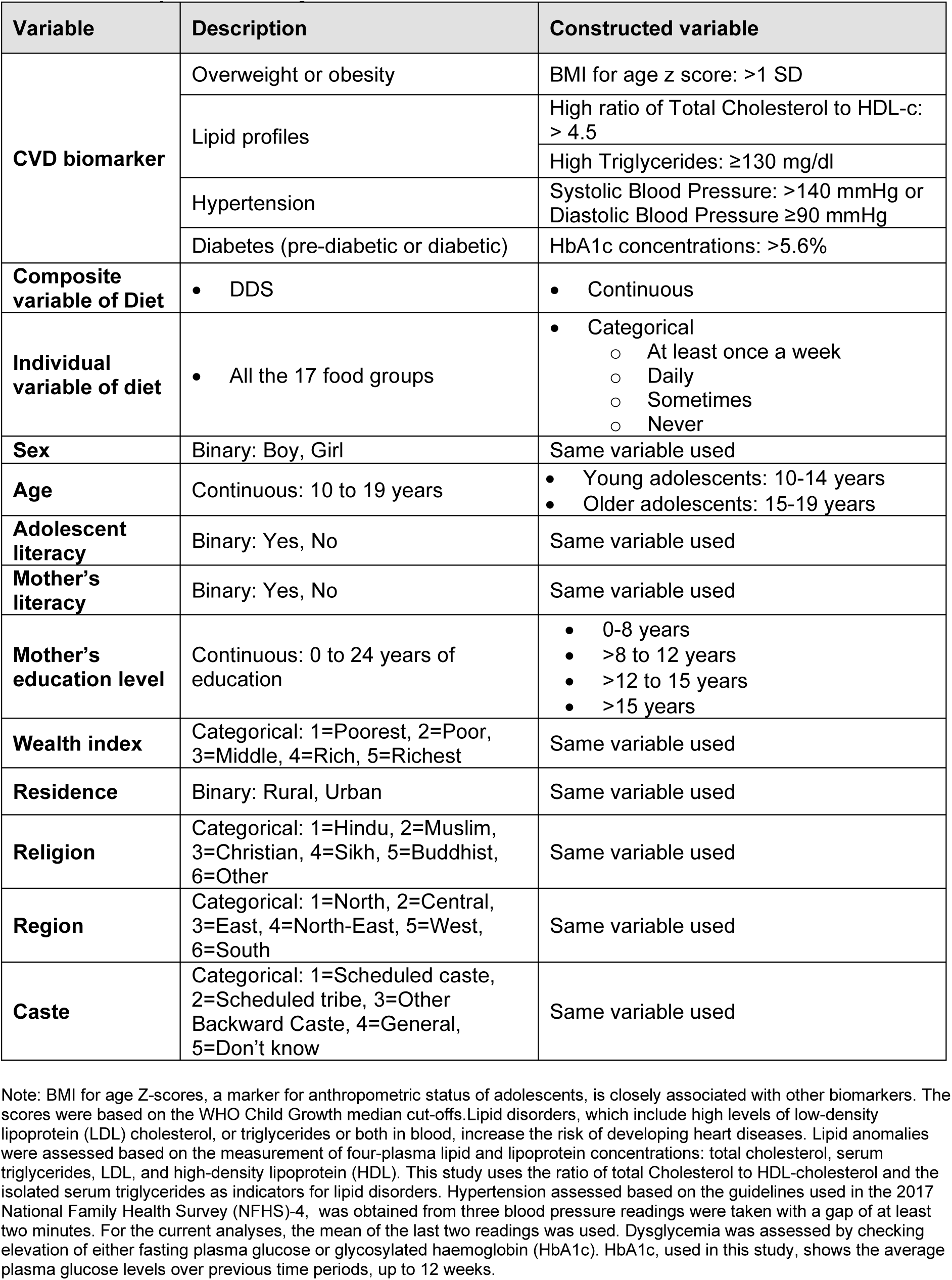
Description of study variables.

### Ethical approval

International and national ethical approvals were obtained from the Population Council’s Institutional Review Board in New York and Post Graduate Institute of Medical Education and Research in Chandigarh, respectively, prior to initiating the CNNS survey. All the respondents underwent an informed consent and assent process for participating in the survey: for 10-year-olds, informed consent was obtained from parent/caregiver; for 11-to-17-year-olds, from both parent/caregivers and adolescents; and for 18-to-19-year-olds, from adolescents. No further ethics committee approval was obtained for this secondary data analysis of the de-identified data.

### Statistical analyses

Analyses were conducted utilising the dietary intake data for all adolescents for whom NCD biomarkers information and socio-economic indicators were available across all 30 Indian states. The survey data were obtained as STATA files. Descriptive analyses were conducted to analyse the sample distribution across a range of background characteristics. T-tests were conducted to examine statistical differences in mean overweight/obesity and the CVD biomarkers by early/late adolescence, gender, and area of residence. Boxplots for overweight/obesity and the CVD biomarkers were plotted to compare the summary statistics by gender, area of residence and early/late adolescence. Bivariate analyses were conducted to understand the dietary intakes and the prevalence of overweight/obesity and the CVD biomarkers by socioeconomic and demographic variables. National weights of biomarker were used in the analyses. STATA version 17.0 was used for the data analysis.

## Results

A breakdown of the characteristics of the sampled population in the CNNS dataset is presented in

**Supplementary Table 1**. Overall, boys and girls were equally represented whereas younger adolescents constituted 52%, rural residents 75%, those attending schools 94%, and Hindus 80%, respectively.

### Dietary pattern

**Table 2 and Figure 1(a)** summarise the mean overall DDS scores across various socio-economic characteristics. Whilst the mean dietary scores are similar between gender and early/late adolescence, they vary significantly by SES (levels of maternal education attainment, household wealth index) and other characteristics including religion, region, and caste. The scores show a positive trend with increase in SES, meaning, adolescents from high SES (i.e., higher levels of - maternal education attainment and -household wealth) have better diet quality.

**Figure 1:**
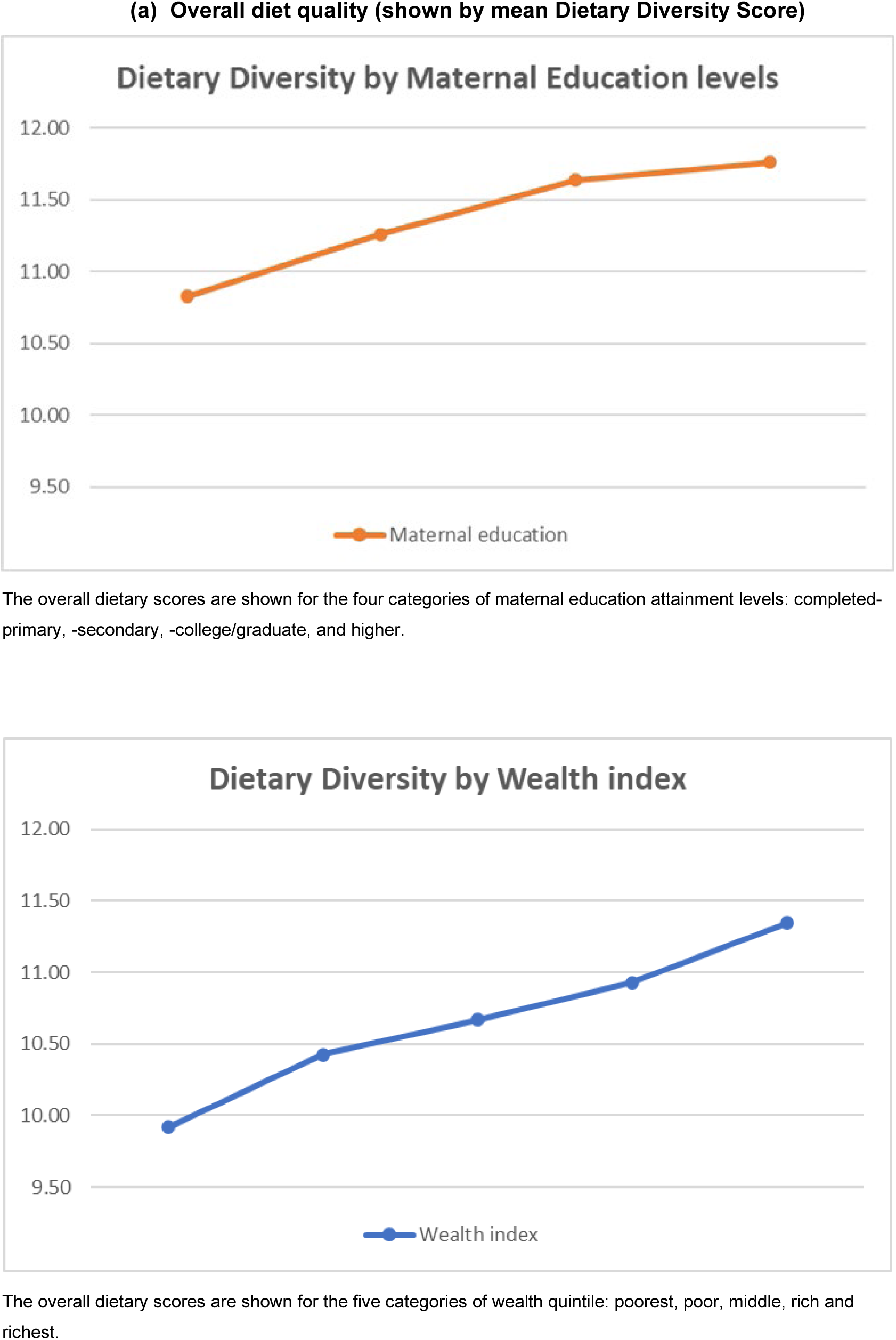
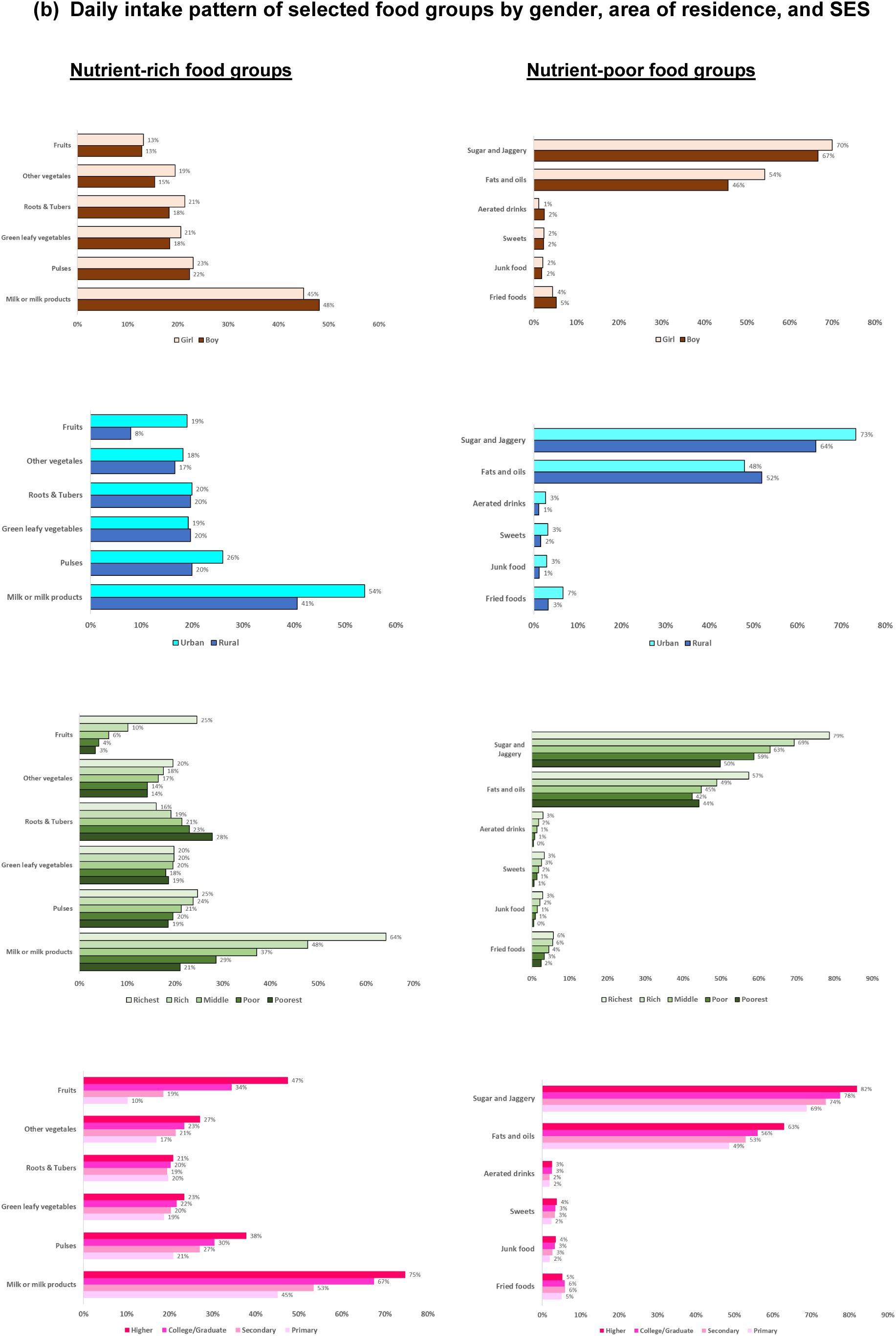
Graphical presentation of dietary pattern among Indian adolescents.

**Table 2.**
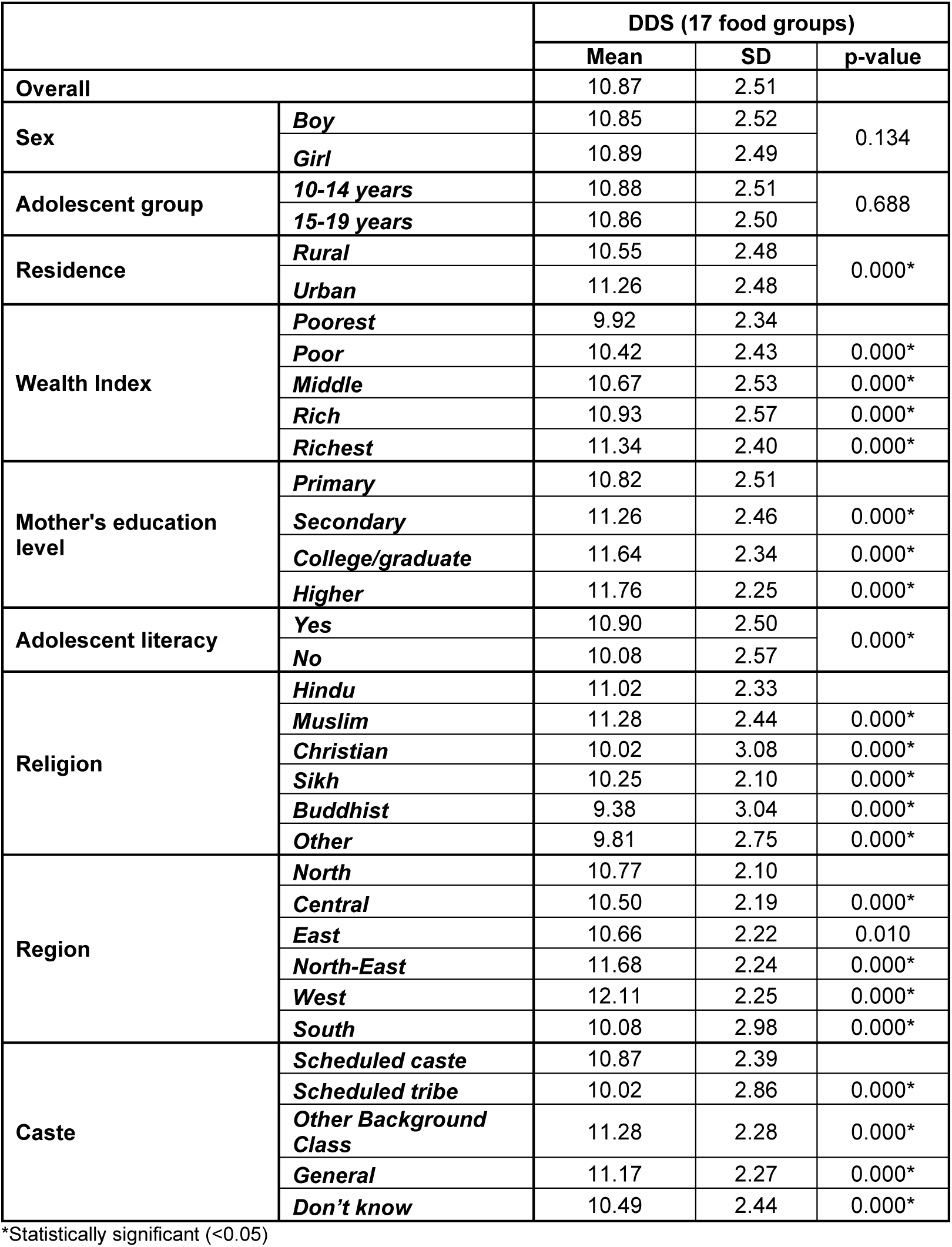
DDS across socio-economic characteristics.

Decomposing the consumption pattern of individual food groups, we found adolescents, in general, have low daily intakes of ICMR^10^ recommended food groups, except for cereals. **[Supplementary Figure 1]** Less than half (47%) and only one-fifth (23%) consume milk or milk products and pulses daily. Daily intakes of fruits and vegetables are at the lowest: only 13% consume fruits daily; 19% green leafy vegetable; 20% roots and tubers; 17% other vegetables, respectively. Frequent intakes (at least once per week) of food items considered non-vegetarian in the Indian context including egg, fish, chicken, or meat ranges between 36-40%. Notably, the daily consumption of fats and oils and sugar and jaggery are very high: 50% of adolescents consume fats and oils and 68% consume sugar and jaggery daily. Majority, ranging between 61%-74%, consume the HFSS food groups occasionally. Breaking down the dietary pattern by gender, area of residence and SES shows daily consumptions of milk and milk products, aerated drinks and fried food are higher among boys than girls. Whereas girls have higher daily consumption of fruits, vegetables, roots and tubers and pulses. Furthermore, higher proportion of adolescents from urban and high SES regularly consume more nutrient-rich foods as well as those high in HFSS, sugar and jaggery, and fats and oils, compared to their counterparts from rural and low SES. See **Figure 1(b)**.

### Prevalence of the CVD biomarkers

Approximately one in four adolescents exhibit abnormalities (meaning, increased risk) in at-least one of the CVD biomarkers: ratio of total cholesterol to HDL-c, isolated serum triglycerides, hypertension, and high levels of-HbA1c.**[Table 3]** Prevalence is higher among girls, urban residents, and in early adolescence.

**Table 3.** Prevalence of any anomalies in CVD biomarkers among Indian adolescents, CNNS 2014-16 (weighted estimates)

|  |  | Any anomalies in CVD biomarkers (95% CI), | p-value |
| --- | --- | --- | --- |
| Overall |  | 26.01% (25.33%, 26.69%) | - |
| Gender | Girl | 26.46% (25.48%, 27.44%) | 0.237 |
|  | Boy | 25.56% (24.62%, 26.50%) |  |
| Area | Urban | 27.02% (25.97%, 28.08%) | 0.000 |
|  | Rural | 25.66% (24.78%, 26.55%) |  |
| Adolescence | 10-14 years | 26.37% (25.44%, 27.30%) | 0.000 |
|  | 15-19 years | 25.60% (24.60%, 26.59%) |  |
| Wealth index | Poorest | 21.23% (19.05%, 23.41%) | - |
|  | Poor | 26.68% (24.83%, 28.54%) | 0.001 |
|  | Middle | 24.54% (23.05%, 26.02%) | 0.000 |
|  | Rich | 26.57% (25.24%, 27.895) | 0.000 |
|  | Richest | 31.02% (29.75%, 32.29%) | 0.000 |
| Maternal education | Literate | 27.75% (26.87%, 28.62%) | 0.000 |
|  | Illiterate | 24.41% (23.32%, 25.49%) |  |
| Levels of maternal education | Primary | 26.44% (25.24%, 27.64%) | - |
|  | Secondary | 30.58% (29.11%, 32.05%) | 0.000 |
|  | College/graduate | 29.80% (26.34%, 33.26%) | 0.000 |
|  | Higher | 25.03% (19.43%, 30.64%) | 0.000 |
| Religion | Hindu | 24.79% (24.01%, 25.57%) | - |
|  | Muslim | 31.8% (29.65%, 33.96%) | 0.266 |
|  | Christian | 33.52% (31.26%, 35.78%) | 0.045 |
|  | Sikh | 29.34% (24.63%, 34.05%) | 0.014 |
|  | Buddhist | 17.82% (13.72%, 21.92%) | 0.024 |
|  | Other | 37.54%(31.31%, 43.78%) | 0.042 |
| Region | North | 24.97% (23.57%, 26.36%) | - |
|  | Central | 21.06% (19.12%, 22.99%) | 0.000 |
|  | East | 29.25% (27.54%, 30.96%) | 0.005 |
|  | North-East | 25.58% (23.58%, 27.59%) | 0.022 |
|  | West | 30.77% (28.92%, 32.63%) | 0.003 |
|  | South | 36.13% (34.59%, 37.66%) | 0.000 |
| Caste | Scheduled caste | 24.76% (23.25%, 26.26%) | - |
|  | Scheduled tribe | 24.5% (22.96%, 26.04%) | 0.406 |
|  | Other background class | 24.89% (23.71%, 26.07%) | 0.669 |
|  | General | 28.54% (27.07%, 30.00%) | 0.000 |
|  | Don't know | 33.12% (28.11%, 38.16%) | 0.925 |

High levels of isolated serum triglycerides are most commonly prevalent among Indian adolescents (16.07%); followed by pre-diabetic/diabetic (9.68%); hypertension (4.99%); overweight/obesity (4.78%); and high ratio of total cholesterol to HDL-c (3.56%) respectively. Overweight/obese is mostly concentrated in urban areas (8.4% versus 3.6% in rural areas) and high SES. Interestingly, the prevalence appears to increase exponentially with improvement in SES. To illustrate, overweight/ obesity is highest among those with higher maternal education (16.92%), which is approximately three times compared to those with primary levels of maternal education attainment (5.90%). Similarly, while the prevalence among poorest quintile is 0.63%, that among the richest quintile is 11.31%, approximately ten times higher. Gender wise, it is marginally higher among boys than girls. For the four remaining biomarkers, there is no clear trend of prevalence by SES. High triglycerides are more prevalent among girls, rural residents, and low SES; pre-diabetes/diabetes is mostly prevalent among boys, rural residents, graduate level of maternal education and richest wealth quintiles. The prevalence of high ratio of total cholesterol to HDL-c and hypertension is overall low among Indian adolescents. **[Table 4 and Figure 2]**

**Figure 2.**
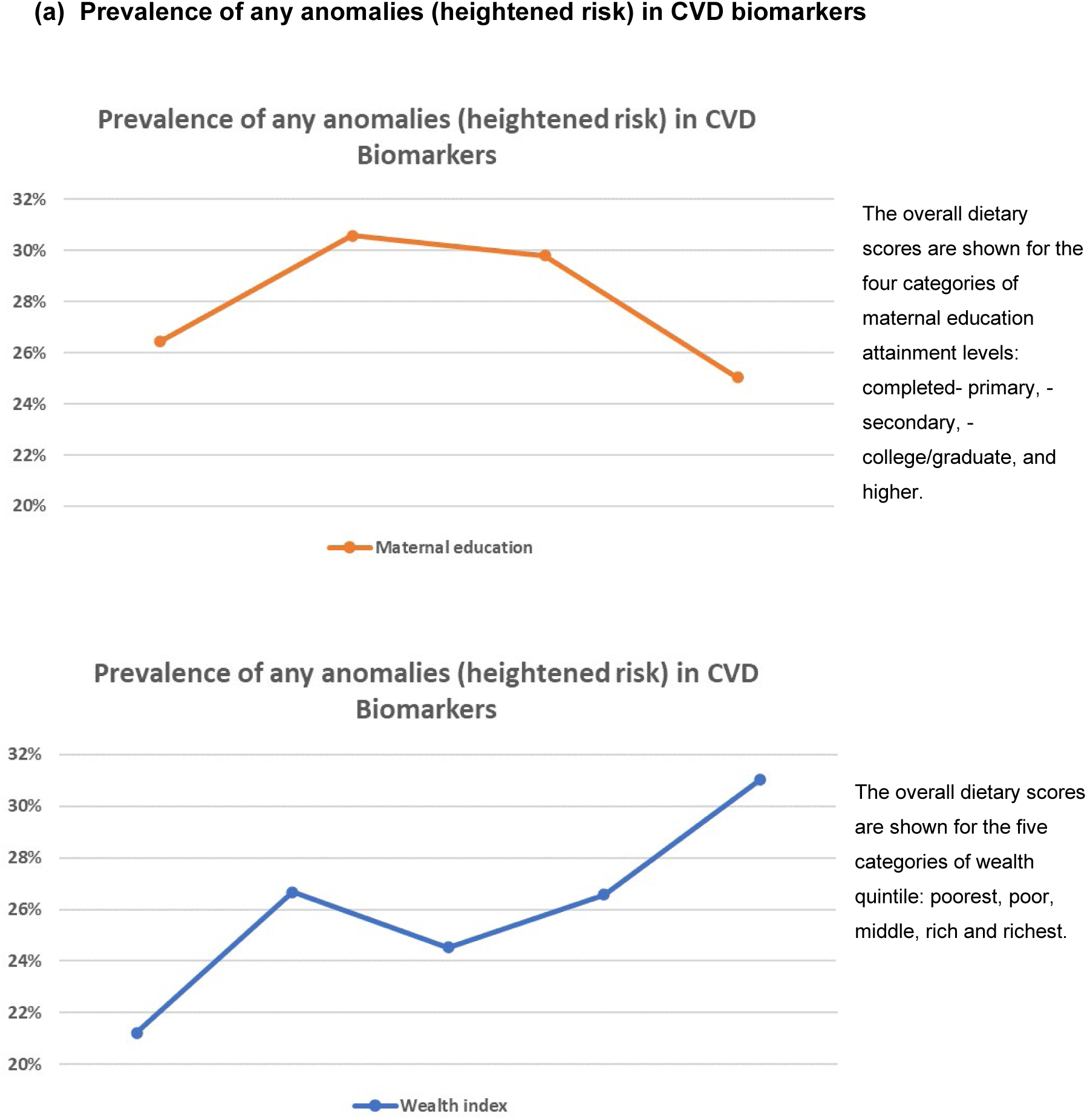
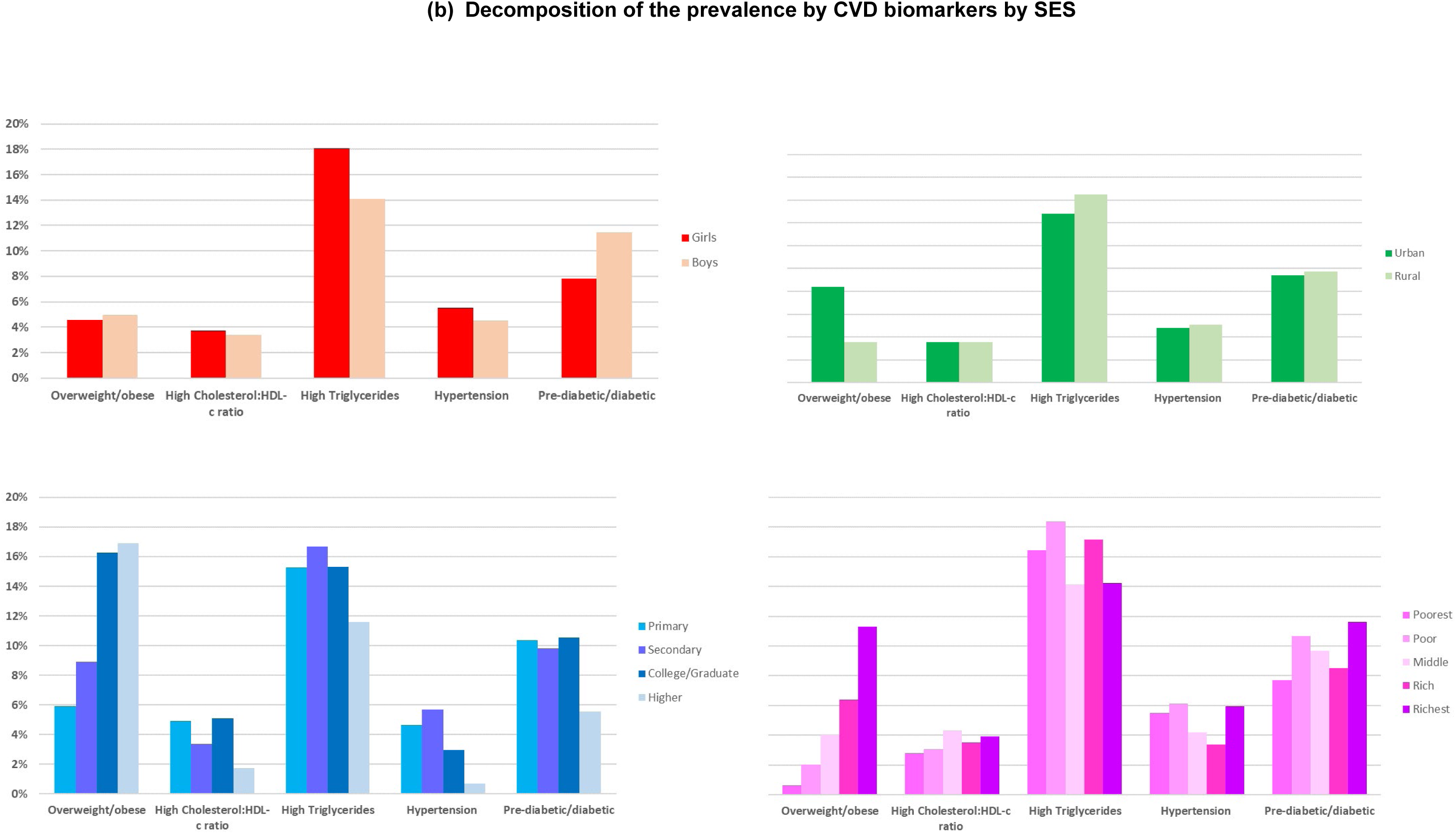
Graphical presentation of CVD prevalence among Indian adolescents by SES.

**Table 4:** Prevalence of CVD biomarkers among Indian adolescents (weighted estimates)

|  |  | BMI-Z score | Lipid anomalies |  | Blood pressure | HbA1c |
| --- | --- | --- | --- | --- | --- | --- |
|  |  | Overweight or obesity | High Total Cholesterol:HD L-c ratio (>4.5) | High Triglycerides | Hypertension (high Systolic or Diastolic BP) | Pre-diabetic or diabetic |
| **Overall** |  | 4.78% | 3.56% | 16.07% | 4.99% | 9.68% |
| **Sex** | *Girls* | 4.58% | 3.73% | 18.08% | 5.51% | 7.85% |
|  | *Boys* | 4.99% | 3.39% | 14.10% | 4.50% | 11.48% |
| **Area of residence** | *Urban* | 8.39% | 3.54% | 14.79% | 4.77% | 9.40% |
|  | *Rural* | 3.58% | 3.57% | 16.50% | 5.07% | 9.77% |
| **Mother's literacy** | *Literate* | 7.40% | 4.36% | 15.65% | 4.84% | 10.10% |
|  | *Illiterate* | 2.37% | 2.79% | 16.47% | 5.14% | 9.28% |
| **Mother's education** | *Primary* | 5.90% | 4.90% | 15.27% | 4.64% | 10.35% |
|  | *Secondary* | 8.92% | 3.39% | 16.66% | 5.70% | 9.81% |
|  | *College/Graduate* | 16.29% | 5.12% | 15.31% | 2.98% | 10.53% |
|  | *Higher* | 16.92% | 1.76% | 11.59% | 0.70% | 5.56% |
| **Wealth** | *Poorest* | 0.63% | 2.81% | 16.44% | 5.52% | 7.72% |
|  | *Poor* | 2.02% | 3.09% | 18.38% | 6.12% | 10.68% |
|  | *Middle* | 4.08% | 4.35% | 14.13% | 4.18% | 9.69% |
|  | *Rich* | 6.39% | 3.51% | 17.14% | 3.39% | 8.53% |
|  | *Richest* | 11.31% | 3.95% | 14.25% | 5.94% | 11.62% |
| **Religion** | *Hindu* | 4.75% | 3.28% | 14.44% | 4.70% | 9.59% |
|  | *Muslim* | 4.37% | 4.63% | 25.63% | 7.22% | 8.72% |
|  | *Christian* | 4.83% | 7.66% | 17.80% | 7.76% | 14.87% |
|  | *Sikh* | 10.99% | 2.85% | 10.00% | 0.64% | 13.65% |
|  | *Buddhist* | 5.43% | 0.28% | 2.20% | 0.18% | 17.12% |
|  | *Other* | 6.18% | 4.88% | 39.31% | 1.98% | 7.14% |
| **Region** | *North* | 4.87% | 2.40% | 12.17% | 2.75% | 13.27% |
|  | *Central* | 1.90% | 2.90% | 14.08% | 6.37% | 7.36% |
|  | *East* | 4.40% | 4.37% | 23.40% | 4.69% | 7.05% |
|  | *North-East* | 7.79% | 4.67% | 10.73% | 4.98% | 13.04% |
|  | *West* | 9.35% | 4.01% | 12.68% | 4.94% | 12.08% |
|  | *South* | 4.92% | 3.03% | 30.91% | 6.00% | 14.83% |
| **Caste** | *Scheduled caste* | 4.13% | 3.02% | 15.84% | 3.86% | 8.58% |
|  | *Scheduled tribe* | 2.19% | 2.82% | 14.97% | 5.40% | 13.90% |
|  | *OBC* | 5.14% | 4.04% | 14.46% | 4.18% | 9.65% |
|  | *General* | 6.72% | 3.45% | 17.07% | 6.29% | 10.00% |
|  | *Don't know* | 2.10% | 4.05% | 23.48% | 12.83% | 7.04% |

National biomarkers weights used.

### Association between unhealthy diet and prevalence of the CVD biomarkers

**Table 5a** presents the proportions of adolescents with any anomalies (meaning, heightened risk) in the CVD biomarkers across the frequency of consumption of unhealthy diet, including fats and oils, sugar and jaggery and HFSS.^27^ Those consuming junk food daily have the highest prevalence of any anomaly (33%); closely followed by those who never had fats and oils (34%) and fried food (33%), respectively. On the other hand, a breakdown of the proportion of adolescents with the CVD biomarkers indicates those with the anomalies have significantly higher daily consumption of two groups-fats & oils and sugar & jaggery, followed by consuming HFSS foods at least once per week or occasionally.**[see Table 5b]** Notably, daily consumption of these two food groups is particularly very high among those from high SES, shown in **Figure 1(b)**.

**Table 5a:** Proportions of adolescents suffering from any anomalies in the CVD biomarkers across the frequency of consumption of unhealthy food including fats and oils, sugar and jaggery and HFSS (weighted estimates)

|  | *At least once per week* | *Daily* | *Occasional* | *Never* |
| --- | --- | --- | --- | --- |
| Fats and oils | 25% | 25% | 26% | 34% |
| Sugar and jaggery | 25% | 25% | 29% | 26% |
| Fried food | 24% | 24% | 27% | 33% |
| Junk food | 25% | 36% | 26% | 27% |
| Sweets | 28% | 29% | 26% | 25% |
| Aerated drink | 27% | 25% | 26% | 26% |

**Table 5b:** Breakdown of the prevalence of the CVD biomarkers across the frequency of consumption of unhealthy food including fats and oils, sugar and jaggery and HFSS (weighted estimates)

|  | BMI z-scores | Lipid anomalies |  | Blood Pressure | HbA1c |
| --- | --- | --- | --- | --- | --- |
|  | *Overweight or obesity* | *High Total Cholesterol:HDL-c ratio (>4.5)* | *High Triglycerides* | *Hypertension (high Systolic or Diastolic BP)* | *Pre-diabetic or diabetic* |
| **<u>Fats & Oils</u>** |  |  |  |  |  |
| at least once per week | 13% | 7% | 10% | 7% | 12% |
| daily | 45% | 45% | 46% | 58% | 54% |
| occasionally | 31% | 37% | 33% | 27% | 26% |
| never | 11% | 11% | 11% | 8% | 9% |
| **<u>Sugar & jaggery</u>** |  |  |  |  |  |
| at least once per week | 9% | 16% | 12% | 12% | 12% |
| daily | 76% | 64% | 62% | 66% | 70% |
| occasionally | 13% | 19% | 24% | 21% | 16% |
| never | 2% | 2% | 2% | 1% | 2% |
| **<u>Fried foods</u>** |  |  |  |  |  |
| at least once per week | 36% | 23% | 26% | 27% | 31% |
| daily | 4% | 1% | 5% | 4% | 3% |
| occasionally | 56% | 69% | 64% | 67% | 62% |
| never | 4% | 6% | 5% | 2% | 4% |
| **<u>Junk food</u>** |  |  |  |  |  |
| at least once per week | 14% | 7% | 9% | 9% | 10% |
| daily | 1% | 0% | 1% | 1% | 1% |
| occasionally | 57% | 61% | 63% | 67% | 58% |
| never | 28% | 32% | 26% | 23% | 30% |
| **<u>Sweets</u>** |  |  |  |  |  |
| at least once per week | 20% | 12% | 14% | 13% | 14% |
| daily | 4% | 2% | 3% | 3% | 2% |
| occasionally | 67% | 75% | 76% | 79% | 77% |
| never | 9% | 11% | 7% | 4% | 7% |
| **<u>Aerated drinks</u>** |  |  |  |  |  |
| at least once per week | 13% | 12% | 9% | 8% | 10% |
| daily | 4% | 2% | 1% | 3% | 2% |
| occasionally | 68% | 74% | 77% | 77% | 76% |
| never | 15% | 12% | 13% | 12% | 13% |

## Discussion

Our study reveals: i) adolescents from higher SES have higher dietary diversity compared to those from lower SES; and ii) one in four adolescents exhibit abnormalities in at-least one of the CVD biomarkers-overweight/obesity, hypertension, high levels of-HbA1c, high ratio of total cholesterol to HDL-c and high isolated serum triglycerides.

The socio-economic patterning of dietary diversity is concentrated on higher daily consumption of certain foods including fats and oil, sugar and jaggery and HFSS, comprising-junk food, fried food, sweets, and aerated drinks. We found a positive SES gradient in the consumption of these food groups: more socio-economically better-off adolescents and those from those from urban areas have higher consumption of these energy-dense, nutrient-poor foods. This trend may explain the concentration of overweight/obesity in this cohort.

Concerning the prevalence of anomalies in the CVD biomarkers, the SES gradient follows an inverted U-shaped for maternal education attainment (better health for the lowest and highest socially advantaged, worse health for those in the middle) whereas, it is positive for household wealth index (worse health for the more economically advantaged). More girls have anomalies compared to boys. This can be attributed to the overall gender disparity in dietary diversity among Indian adolescents.^27 43^ Similarly, there are significant differences between urban and rural residents.^27 28^ Comparing the prevalence of these anomalies across the intakes of unhealthy food groups, adolescents consuming fats & oils, sugar & jaggery, daily and those consuming HFSS foods at least once per week or occasionally are more likely to have higher prevalence of anomalies in the CVD biomarkers. All these factors together indicate a serious health burden waiting to unfold in the Indian healthcare system.^27 44-48^

A generic narrative of socio-economic patterning of CVD biomarkers in India is problematic as it varies by individual SES variables. To illustrate, the SES gradient for maternal education is reversing in India. However, our analyses, consistent with previous studies,^18 19 32^ showed a clustering of the anomalies in CVD biomarkers among those from higher wealth quintiles. This finding contradicts the narrative that the association between SES and prevalence of CVD biomarkers/ CVD burden has reversed to being negative in India, as shown by Wetzel et al.^15^ The difference in the results could potentially be explained by the differences across the two studies. Firstly, Wetzel et al. examined the prevalence of tobacco consumption, unhealthy weight, diabetes, and hypertension as CVD biomarkers. Secondly, they included different population age groups (15-45 years) and used different data source-National Family Health Survey Rounds 4 and 5. Finally, they measured the change in the prevalence overtime whereas our findings are based on cross-sectional data.

Our findings are consistent with the empirical literature from adult populations suggesting CVD biomarkers are commonly prevalent among the higher SES groups.^18 19^ However, there appears to be a bias towards a particular interpretation that CVD and its biomarkers disproportionately affect the poor in India. While a disproportionate focus may risk shifting the limited healthcare resources from addressing the health concerns of the poor to those of the middle-class and rich in India, it should be acknowledged that the most important inequality disadvantaging the poor is with respect to treatment, rather than incidence of the burden.^49^ Finally, the association between SES and CVD from the lens of Western ‘reversal’ in SES gradient may not be applicable in the Indian context due to the macro socio-economic realities among the Indian population.

This study uses the CNNS dataset which provides data on an understudied age group. No other nation-wide surveys, except NFHS-5, have included data on adolescent population, although the NFHS-5 includes data on late adolescence, 15-19 years. No previous nation-wide survey included data on early adolescence, 10–14-year-olds. The survey not only provides large comprehensive data on nutritional status of Indian children, aged 0-19 years, but also used gold standard methods to assess NCD biomarkers in India.

This study has limitations, which merit consideration. First, the CNNS is a cross-sectional survey, therefore no inference can be made on the shift of dietary behaviour and prevalence of anomalies in the CVD biomarkers overtime. Secondly, data on some of the inter-related factors with diet such as physical activity are not available for examination. Thirdly, data on dietary intake only included frequency of consumption of food groups; a detailed dietary assessment is necessary to conduct an in-depth analysis of diet quality and its potential contributions to CVD biomarkers. Finally, the CNNS survey did not include a measure of household expenditure. In present-day India, the household wealth index may be an unreliable indicator of household income within urban areas, as most households possess majority of the household assets included to measure this index.

In conclusion, there is a clustering of the prevalence of anomalies in the CVD biomarkers among adolescents from urban and wealthier households, which may be associated with an increased consumption of energy-dense, nutrition-poor food groups in this population. This warrants immediate, active, and appropriate diet related and broader lifestyle interventions targeting this cohort.^26^

## Data Availability

The Comprehensive National Nutrition Survey Data team provided all data produced in the present work. All data produced in the present work are contained in the manuscript.

## Acknowledgements

The authors thank Professor Richard Cookson, Dr Wiktoria Tafesse and Dr Claire Carswell for commenting on an earlier version of the draft.

## Author contributions

NK conceived the idea, conducted all the statistical analyses, and drafted the manuscript. SG and SM guided the analysis and provided analytical insight for the CNNS data. All authors were involved at each iteration of drafting and approved the final paper. All authors had access to raw data.

## Data Sharing

The Ministry of Health and Family Welfare (MoHFW), Government of India owns the Comprehensive National Nutrition Survey data. The data used in this paper were released for public use by the MoHFW and United Nations Children Fund, India Country Office. The analytic code for the analyses can be made available upon request to the corresponding author(s).

## Compliance with ethical standards

## Ethical approval

The Comprehensive National Nutrition Survey was conducted according to the guidelines laid down in the Declaration of Helsinki and all procedures involving human subjects were approved by the Population Council’s International Review Board (New York, USA) and ethics committee of Post Graduate Institute of Medical Education and Research (Chandigarh, India). No separate ethical approval was required or taken for this secondary analysis.

