## Supplementary Table 1 for "Socio-economic patterns of diet, obesity, and biomarkers for cardiovascular disease among Indian adolescents"

**Supplementary Document**

Table 1 Sociodemographic characteristics of adolescents aged 10-19 years (weighted)

| **Characteristic** | | **10-14 years** | | | | **15-19 years** | | | | **Overall (10-19 years)** | | | | | **Total** | | | | **Cumulative total** |
| --- | --- | --- | --- | --- | --- | --- | --- | --- | --- | --- | --- | --- | --- | --- | --- | --- | --- | --- | --- |
|  |  | **Boy** | | **Girl** | | **Boy** | | **Girl** | | **Boy** | | **Girl** | | |  |  |  |  |  |
|  |  | **N** | **%** | **N** | **%** | **N** | **%** | **N** | **%** | **N** | **%** | **N** | **%** | **N** | | **%** | |  | |
| ***Sex*** |  | 9,460 | 26% | 9,116 | 25% | 8,404 | 23% | 8,849 | 25% | 17,865 | 50% | 17,965 | 50% |  | |  | | 35,830 | |
| ***Attended school*** | Yes | 9,374 | 26% | 8,428 | 24% | 8,166 | 23% | 7,795 | 22% | 17,540 | 49% | 16,223 | 45% | 33,763 | | 94% | |  | |
|  | No | 383 | 1% | 403 | 1% | 502 | 1% | 779 | 2% | 885 | 2% | 1182 | 3% | 2,067 | | 6% | | 35,830 | |
| ***Religion*** | Hindu | 7,892 | 22% | 6991 | 20% | 7067 | 20% | 6811 | 19% | 14,959 | 42% | 13,802 | 39% | 28,762 | | 80% | |  | |
|  | Muslim | 1419 | 4% | 1449 | 4% | 1185 | 3% | 1387 | 4% | 2,604 | 7% | 2,837 | 8% | 5,441 | | 15% | |  | |
|  | Christian | 239 | 1% | 191 | 1% | 209 | 1% | 179 | 1% | 448 | 1% | 370 | 1% | 818 | | 2% | |  | |
|  | Sikh | 119 | 0% | 98 | 0% | 109 | 0% | 100 | 0% | 228 | 1% | 198 | 1% | 427 | | 1% | |  | |
|  | Buddhist | 50 | 0% | 52 | 0% | 72 | 0% | 59 | 0% | 123 | 0% | 111 | 0% | 234 | | 1% | |  | |
|  | Other* | 37 | 0% | 50 | 0% | 25 | 0% | 37 | 0% | 63 | 0% | 87 | 0% | 149 | | 0% | | 35,830 | |
| ***Caste*** | Scheduled caste | 2188 | 6% | 1861 | 5% | 1903 | 6% | 1876 | 6% | 4091 | 12% | 3737 | 11% | 7828 | | 23% | |  | |
|  | Scheduled tribe | 1097 | 3% | 891 | 3% | 821 | 2% | 963 | 3% | 1917 | 6% | 1854 | 5% | 3771 | | 11% | |  | |
|  | Other Backward Caste | 3766 | 11% | 3620 | 11% | 3572 | 11% | 3518 | 10% | 7338 | 22% | 7138 | 21% | 14477 | | 43% | |  | |
|  | General | 1791 | 5% | 1594 | 5% | 1693 | 5% | 1561 | 5% | 3484 | 10% | 3154 | 9% | 6638 | | 20% | |  | |
|  | Don’t know | 409 | 1% | 423 | 1% | 217 | 1% | 243 | 1% | 626 | 2% | 665 | 2% | 1291 | | 4% | | 34,006 | |
| ***Residence*** | Rural | 7422 | 21% | 6716 | 19% | 6366 | 18% | 6458 | 18% | 13,788 | 38% | 13,174 | 37% | 26,962 | | 75% | |  | |
|  | Urban | 2335 | 7% | 2115 | 6% | 2302 | 6% | 2115 | 6% | 4,637 | 13% | 4,231 | 12% | 8,868 | | 25% | | 35,830 | |
| ***Region*** | North | 1665 | 5% | 1314 | 4% | 1361 | 4% | 1320 | 4% | 3027 | 8% | 2633.9 | 7% | 5661 | | 16% | |  | |
|  | Central | 2551 | 7% | 2158 | 6% | 2107 | 6% | 2974 | 8% | 4658 | 13% | 5131.9 | 14% | 9790 | | 27% | |  | |
|  | East | 2680 | 7% | 2629 | 7% | 1711 | 5% | 1842 | 5% | 4391 | 12% | 4471.6 | 12% | 8863 | | 25% | |  | |
|  | North-East | 1091 | 3% | 996 | 3% | 1051 | 3% | 735 | 2% | 2141 | 6% | 1731 | 5% | 3872 | | 11% | |  | |
|  | West | 1053 | 3% | 1048 | 3% | 1152 | 3% | 870 | 2% | 2205 | 6% | 1918.3 | 5% | 4124 | | 12% | |  | |
|  | South | 1229 | 3% | 825 | 2% | 773 | 2% | 693 | 2% | 2002 | 6% | 1518.4 | 4% | 3520 | | 10% | | 35830 | |
| ***Wealth index*** | Poorest | 2187 | 6% | 1871 | 5% | 1429 | 4% | 1679 | 5% | 3616 | 10% | 3550 | 10% | 7166 | | 20% | |  | |
|  | Poor | 2001 | 6% | 1876 | 5% | 1674 | 5% | 1610 | 4% | 3676 | 10% | 3486 | 10% | 7162 | | 20% | |  | |
|  | Middle | 1859 | 5% | 1719 | 5% | 1837 | 5% | 1754 | 5% | 3696 | 10% | 3473 | 10% | 7169 | | 20% | |  | |
|  | Rich | 1729 | 5% | 1746 | 5% | 1885 | 5% | 1799 | 5% | 3614 | 10% | 3545 | 10% | 7158 | | 20% | |  | |
|  | Richest | 1981 | 6% | 1619 | 5% | 1843 | 5% | 1733 | 5% | 3824 | 11% | 3351 | 9% | 7175 | | 20% | | 35830 | |
| ***Mother's education*** | Literate | 4937 | 14% | 4328 | 12% | 4012 | 11% | 3630 | 10% | 8949 | 25% | 7958 | 22% | 16,907 | | 47% | |  | |
|  | Illiterate | 4820 | 13% | 4504 | 13% | 4656 | 13% | 4943 | 14% | 9476 | 26% | 9447 | 26% | 18,923 | | 53% | | 35,830 | |
| ***Mother's level of education*** | Primary (upto 8 years) | 3619 | 16% | 3312 | 15% | 3070 | 14% | 2999 | 14% | 6690 | 30% | 6311 | 28% | 13,001 | | 59% | |  | |
|  | Secondary (upto 12 years) | 2203 | 10% | 2026 | 9% | 1795 | 8% | 1588 | 7% | 3998 | 18% | 3614 | 16% | 7612 | | 34% | |  | |
|  | Graduate (upto 15 years) | 306 | 1% | 342 | 2% | 215 | 1% | 213 | 1% | 521 | 2% | 555 | 2% | 1076 | | 5% | |  | |
|  | Higher (≥16 years) | 180 | 1% | 165 | 1% | 87 | 0% | 89 | 0% | 267 | 1% | 255 | 1% | 522 | | 2% | | 22,211 | |
| ***Source: CNNS dataset*** | |  |  |  |  |  |  |  |  |  |  |  |  |  | |  | |  | |
| *Other religions include Jain, Jewish, Parsi/Zoroastrian, no religion, and other non-denominated religions | | | | | | | | | |  |  |  |  | |  | |  | |  |

**Figure 1: A breakdown of the consumption pattern of 17 different food groups included in the CNNS dataset for adolescents aged 10-19 years**

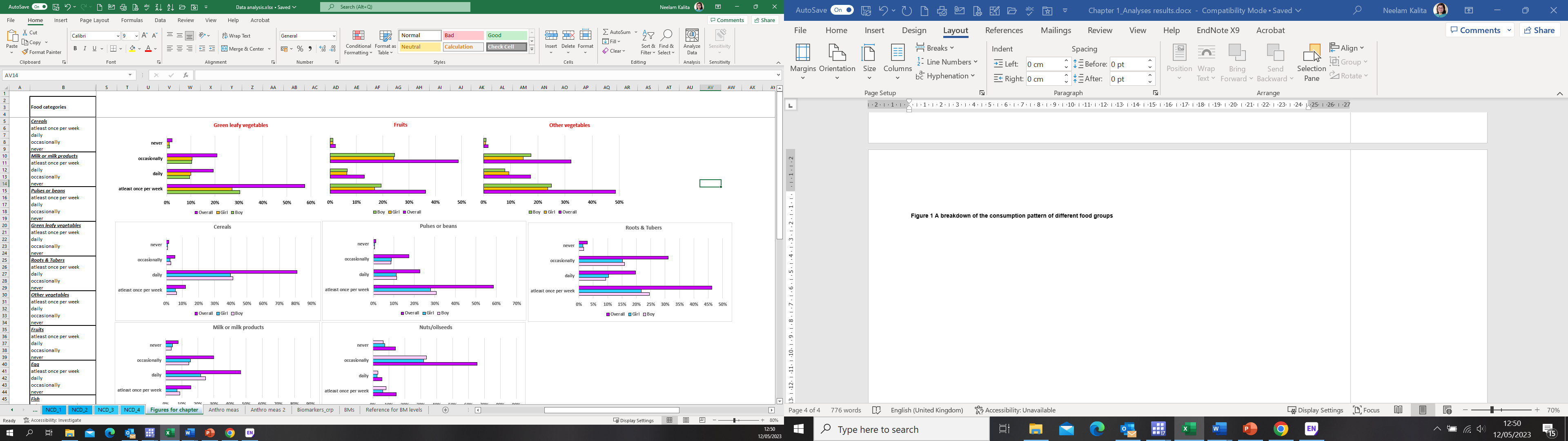

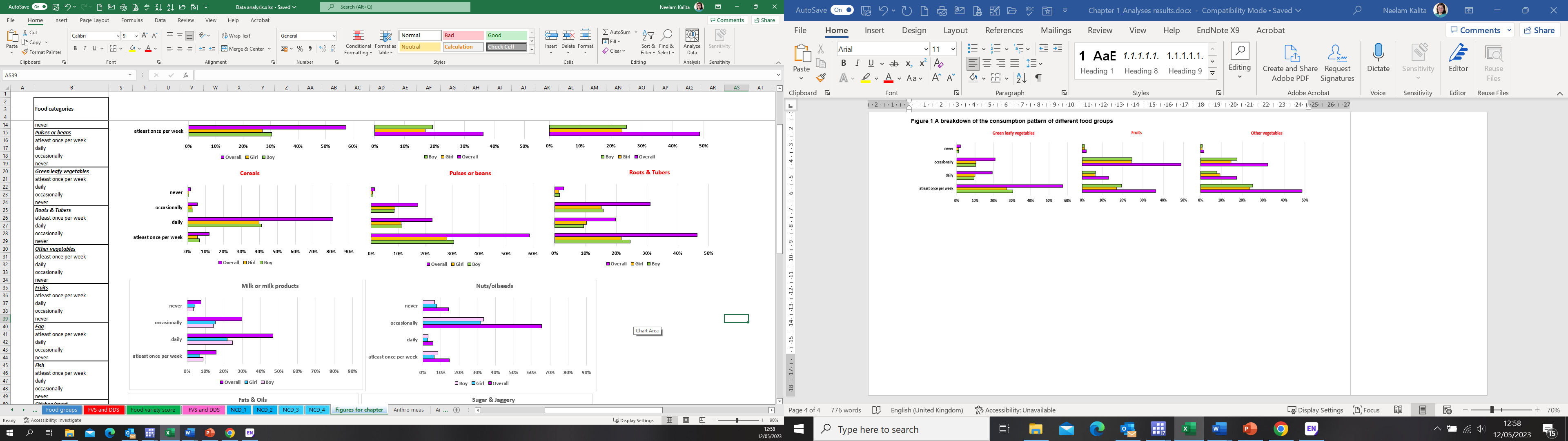

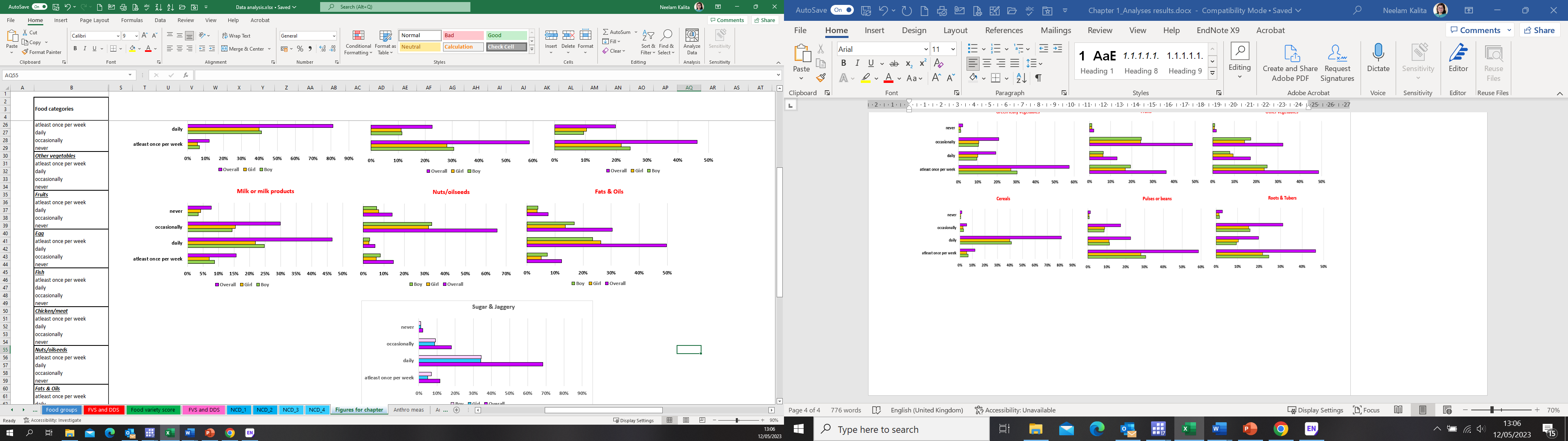

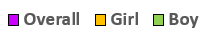

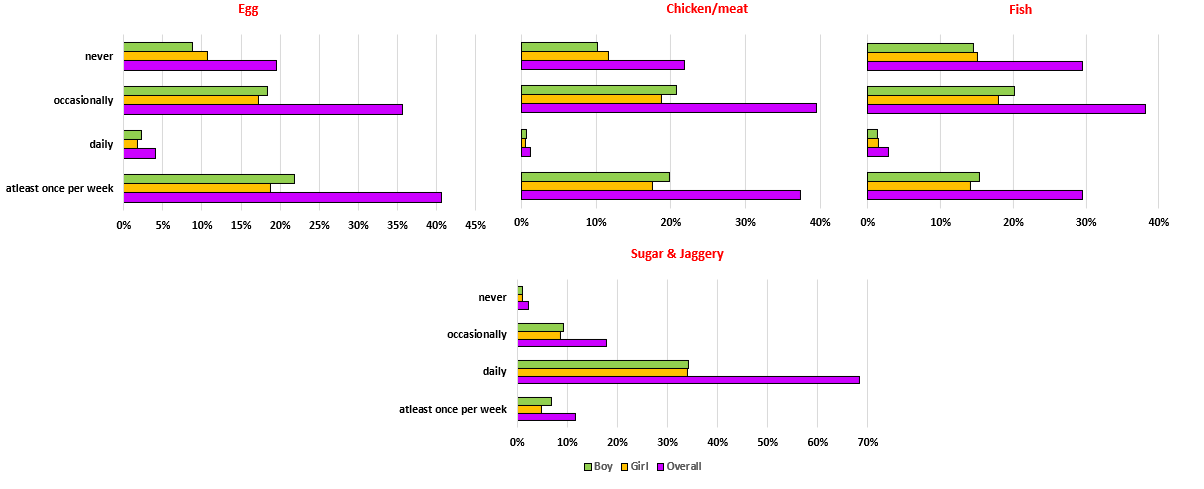

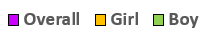

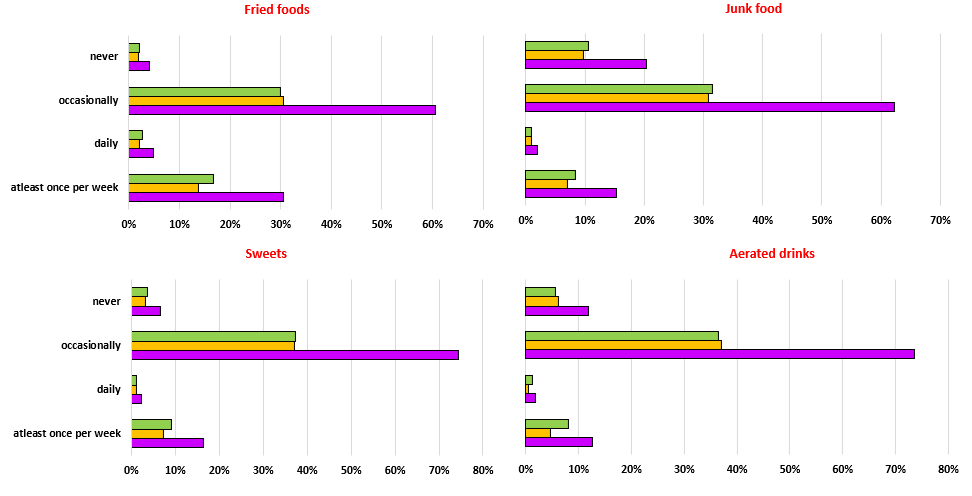

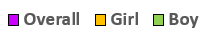
